# Effect of prior tuberculosis on cardiovascular status in perinatally HIV-1-infected adolescents

**DOI:** 10.1101/2024.03.09.24303989

**Authors:** Itai M Magodoro, Carlos E Guerrero-Chalela, Landon Myer, Jennifer Jao, Mpiko Ntsekhe, Katalin A Wilkinson, Robert J Wilkinson, Heather Zar, Ntobeko AB Ntusi

## Abstract

Whether, and how, co-occurring HIV-1 infection (HIV) and tuberculosis (TB) impact cardiovascular status, especially in adolescents with perinatally acquired HIV (APHIV), have not been examined. We hypothesized that APHIV with previous active TB have worse cardiac efficiency than APHIV without TB, which is mediated by increased inflammation. Arterial elastance (Ea) and ventricular end-systolic elastance (Ees) were assessed by cardiovascular magnetic resonance, and ventriculoarterial coupling (VAC) estimated as Ea/Ees ratio. Inflammation was measured by high sensitivity C-reactive protein (hsCRP). Previous TB in APHIV was associated with reduced cardiac efficiency, related to an altered ventriculoarterial coupling. However, we did not find evidence of hsCRP mediated effects in the association between prior TB and cardiac efficiency. The clinical significance of these findings requires further study, including a wider range of biomarkers of specific immune pathways.

## Introduction

Chronic or prolonged infections have been linked with heightened risk of cardiovascular disease (CVD).^1, 2^ Chronic infection-induced cellular and humoral immune activation and systemic inflammation may contribute to endothelial dysfunction, hypercoagulability, atherosclerosis, myocardial inflammation and fibrosis.^3, 4^ These pathological changes precede and characterize adverse cardiovascular events in the general population. In this regard, HIV-1 is now well recognized as an independent risk factor for CVD.^5, 6^ Lately, research attention has been drawn to likely long-term detrimental cardiovascular effects of infection with *Mycobacterium tuberculosis (M.tb).*^7, 8^ A recent (2020) meta-analysis summarizing extant studies, estimated the risk of incident CVD with prior tuberculosis (TB) to be 1.5 times [risk ratio 1.51 (95%CI: 1.16-1.97)] higher than without a history of TB.^9^ Of note is that the included studies were largely drawn from high-income countries (HICs). If TB does prove to be a risk factor for CVD, the challenge will be immense in sub-Saharan Africa (SSA) where endemic *M.tb* coincides with pandemic HIV, pervasive traditional risk factors like hypertension and cigarette smoking, and scant health resources.^10, 11^

However, there is a dearth of studies in SSA examining cardiovascular health in HIV/TB comorbidity, and especially among vulnerable population groups like those with perinatally acquired HIV-1 (PHIV). Children with PHIV are increasingly reaching adolescence and early adulthood due to successful antiretroviral treatment (ART). While many are thriving, a significant proportion face unprecedented multi-system and multi-organ morbidity, including the cardiovascular system, with prospects for poor long-term outcomes.^12, 13, 14^ Adolescents with PHIV (APHIV) are exposed to HIV and ART related cardiotoxicity beginning in infancy, if not *in utero*^15, 16^; and notwithstanding ART, have up to fourfold increased risk of developing TB.^17^ In South Africa, HIV, TB, cerebrovascular disease and CVD are leading causes of premature adult mortality.^18, 19^ Their multimorbidity^20, 21^ is very common, raising the possibility that immune activation and systemic inflammation associated with co-occuring HIV/TB likely negatively impacts the cardiovascular system.^7, 8^ APHIV experiencing TB may therefore be predisposed to premature cardiac morbidity and mortality as they enter adulthood.

Assessment of ventricular-arterial coupling (VAC) provides important insights into the pathophysiology of heart failure including its preclinical antecedents.^22^ VAC simultaneously evaluates ventricular performance and arterial hemodynamics, and the degree of matching between the two. Optimal VAC allows the heart to effectively pump blood into the arterial circulation while minimizing excess workload.^22, 23, 24^ Mismatched coupling, where arterial load and ventricular contractility do not correspond, can lead to inefficient cardiac function and potential heart failure. VAC is therefore an integrative characterization of cardiovascular system dynamics.^22^ Combined with assessment of systemic immune changes, evaluation of VAC in APHIV may yield important, actionable insights into early cardiac disease and its pathophysiology. In this cross-sectional study, we aimed to evaluate the effect of TB infection on cardiac status of adolescents growing up with PHIV in Cape Town, South Africa. We hypothesized that APHIV with previous TB will have worse VAC than peers without, because of synergism of TB/HIV on immune activation and systemic inflammation.

## Methods

### Study participants

Participants were drawn from an ongoing longitudinal (parent) study of chronic diseases development in Cape Town.^25^ Cohort members include APHIV and their age-, sex- and community-matched HIV uninfected peers. All APHIV participants were stably on ART, initiated in childhood. For the present analysis, APHIV presenting for a scheduled visit in the parent study were consecutively approached with invitation to undergo cardiovascular magnetic resonance (CMR) examination in addition to routine study procedures. All were eligible for inclusion if they had no current cardiorespiratory symptoms or known structural heart disease, active systemic infection, and had no contra-indications to CMR.

### Tuberculosis disease

We ascertained TB status from electronic medical records. We defined having previous TB as any history of a clinician-led diagnosis of TB, prescription of standard antituberculosis drug regimen, and/or positive GeneXpert MTB/RIF and/or microscopy and culture of acid-fast bacilli prior to CMR examination. Data on TB site and treatment completion were lacking, while data on dates when TB was diagnosed were incomplete to allow us to estimate time intervals between TB episode and CMR examination.

### CMR image acquisition and analysis

The CMR protocol was similar to that previously published by the present authors.^26^ Briefly, we performed all scans at 3 Tesla on a Siemens Skyra MR system (Erlangen, Germany), and acquired long axis (LAX) and short axis (SAX) cines using the breath-hold steady-state free precession (bSSFP) sequence. Exams also included T2-weighted imaging for assessment of myocardial edema and native T1 mapping for diffuse fibrosis. Late-gadolinium enhancement (LGE) images were acquired to assess for scar/fibrosis and post-contrast T1 mapping was performed to calculate extracellular volume (ECV). Image analysis^27^ was completed offline blinded to PHIV status and using the proprietary CVI42^®^ (Circle Cardiovascular Imaging, Calgary, Canada). Endo- and epicardial contours were automatically generated with manual correction where warranted. LV end-diastolic volume (LVEDV), LV ejection fraction (LVEF) and LV mass (LVM) were calculated on SAX cines. All volumetric and mass data were indexed to height^1.7^ and this is indicated by postscript (i).

Strain analysis was done on the SAX and LAX (2Ch, 3Ch, and 4Ch) cines using automatic feature tracking. We extracted global peak systolic circumferential (GCS) and longitudinal (GLS) strain, and global peak diastolic circumferential and longitudinal strain rates. Motion corrected T1, T2 and extracellular volume (ECV) maps were generated using basal, mid-ventricle, and apical SAX slices. Global measurements per slice were averaged to yield mean native T1, native T2 and ECV values, while LGE was visually scored for presence (yes/no). We calculated the VAC ratio (the primary endpoint) as the ratio of arterial elastance (Ea) to ventricular end-systolic elastance (Ees), where Ea = mean arterial blood pressure/ventricular stroke volume and Ees = mean arterial blood pressure/end-systolic volume.^23, 28, 29, 30^ Ea represents the load on the heart, while Ees signifies ventricular contractility.

### HIV markers and antiretroviral treatment history

Participants had their CD4+ cell count and HIV viral load (detection limit <40 RNA copies/mL, Roche Cobas AmpliPrep/TaqMan, Pleasanton, CA, USA) measured at the time of CMR examination. Data on current and past ART were retrieved from electronic medical records.

### High sensitivity C-reactive protein and cardiometabolic markers

The Tina-quant CRPHS immunoturbidimetric assay was used for the quantitative determination of high sensitivity C-reactive protein (hs-CRP). Participants were excluded from analysis if hs-CRP >10 mg/dL as this could signify an additional infection or an inflammatory condition.^31^ The last two of three attended brachial blood pressure (BP) measurements taken using electronic sphygmomanometers were averaged, and mean arterial pressure (MAP) determined. Z scores for body mass index (BMI) was calculated from weight (kg) and height (m) and compared to the WHO reference population^32^ to obtain BMI z scores.

### Ethics approval

The Human Research Ethics Committee (HREC) of the Faculty of Health Sciences of the University of Cape Town (HREC 051–2013) approved all study activities. Signed informed consent if ≥16 years old or informed parental signed consent and participant assent if <16 years old was obtained prior to participation in the study.

### Data availability

Data used in this study are available upon reasonable request to the authors.

## Data analysis

To test our hypothesis that PHIV with TB is associated with worse VAC than PHIV alone, and that this is driven by increased inflammation/immune activation (measured by hsCRP), we planned *a priori* a causal mediation analysis using the counterfactual framework. A mediator is defined as a variable that is on the causal pathway between the exposure and outcome of interest (**Supplementary Figure 1**). Thus, performing mediation analysis allows the identification of pathways by which an exposure impacts an outcome. Formally, mediation is present if the following four conditions are met: (1) the exposure is associated with the outcome of interest; (2) the exposure is associated with the potential mediator; (3) the potential mediator is associated with the outcome of interest; and (4) including both exposure and mediator as predictors changes the magnitude of association between the outcome of interest and exposure in condition (1).^33^

With past TB infection (yes/no) as the exposure of interest, we built linear regression models of VAC indices to test condition (1), and quintile regression models of hsCRP to test condition (2). To test condition (3), we linearly regressed VAC indices on hsCRP. All regression models were adjusted for age, sex, BMI, HIV viral suppression, and exposure to protease inhibitors (PI) and nonnucleoside reverse transcriptase inhibitors (NNRTI). However, conditions (2) and (3) were not met (*as outlined in **Results** section*), i.e., there was no evidence of mediation by hsCRP, and we therefore did not proceed to test condition (4). The remainder of the analyses focused on history of TB (exposure) and its association with VAC indices using traditional statistical approaches.

Analyses were conducted using R, version 3.6.3 (R Foundation for Statistical Computing, Vienna, Austria), and Stata version 17.0 (StataCorp, College Station, TX, USA). All probability values were 2-sided, with p-values <0.05 considered indicative of statistical significance. Participants’ demographic, HIV and cardiometabolic characteristics were summarized by TB status. Depending on variable scale, we reported values as mean (SD) or median (IQR) or number (%). Differences in cardiac indices and hsCRP according to TB status were examined using linear and quintile regression models, respectively. The β-coefficients from the latter were reported as mean difference in median value. Adjustment was made for age, sex, BMI, HIV viral suppression, and exposure to protease inhibitors (PI) and nonnucleoside reverse transcriptase inhibitors (NNRTI).

## Results

### Cohort description

We enrolled 70 APHIV, 43 of whom had previous TB disease and 27 did not (**Table 1**). The two exposure groups had comparable age and sex composition. They also had similar age at ART initiation and total duration of ART exposure. However, their experience with specific ART regimen was different, as were their clinical outcomes. For example, APHIV without TB had longer median (IQR) NNRTI exposure [7.6 (4.7, 11.0) years)] than their peers with TB [4.2 (0.5, 8.6) years]. Conversely, those with TB had more frequent viral suppression (67%) than APHIV without TB (58%).

**Table 1.** Characteristics of perinatally HIV-1-infected adolescents according to tuberculosis disease status.

| Characteristic | Clinical tuberculosis disease status |  |
| --- | --- | --- |
|  | No prior TB | Prior TB |
| Number | 27 | 43 |
| Age (years, S.D.) | 15.4 (1.7) | 15.0 (1.5) |
| Male sex | 16 (59%) | 22 (51%) |
| ***HIV infection and ART*** |  |  |
| Age at HAART initiation (years) | 3.6 (0.9, 6.7) | 3.3 (1.5, 5.5) |
| Lifetime ART exposure (years) | 11.4 (9.3, 13.6) | 11.5 (10.1, 13.4) |
| NRTI exposure (years) | 11.4 (9.3, 13.6) | 11.5 (10.1, 13.4) |
| PI exposure (years) | 0.6 (0.0, 5.6) | 5.9 (0.0, 10.3) |
| NNRTI exposure (years) | 7.6 (4.7, 11.0) | 4.2 (0.5, 8.6) |
| Current CD4+ count (cells/mL) | 651 (518, 848) | 774 (527, 978) |
| Undetectable HIV viral load | 15 (58%) | 28 (67%) |
| ***Anthropometry*** |  |  |
| BMI (kg/m<sup>2</sup>) | 20.6 (3.8) | 21.3 (5.7) |
| BMI-for-age z-score | 0.00 (1.04) | 0.08 (1.49) |
| Height-for-age z-score | -0.88 (1.25) | -1.03 (1.03) |
| Weight-for-age z-score | -0.44 (1.22) | -0.37 (1.49) |
| ***Blood pressure*** |  |  |
| Mean arterial BP (mmHg) | 113.2 (7.5) | 111.5 (10.2) |
| Systolic BP (mmHg) | 104.6 (7.2) | 103.3 (9.7) |
| Diastolic BP (mmHg) | 65.0 (5.1) | 64.1 (6.5) |

Values are reported as mean (SD) or median (25<sup>th</sup>, 75<sup>th</sup> percentile) or number (%).

### High sensitivity C-reactive protein

There were no statistically significant differences in hsCRP between APHIV with previous TB [median (IQR): 1.53 (0.53, 11.8) mg/dL] and their peers without TB [0.98 (0.40, 2.57) mg/dL; p = 0.29] (**Figure 1** and **Table 2**). When adjusting for potential confounders, having unsuppressed HIV was the only significant predictor of hsCRP: viremia was associated with a 9.78 (0.50, 19.1) mg/dL (p = 0.039) higher median hsCRP compared to undetectable virus.

**Figure 1.**
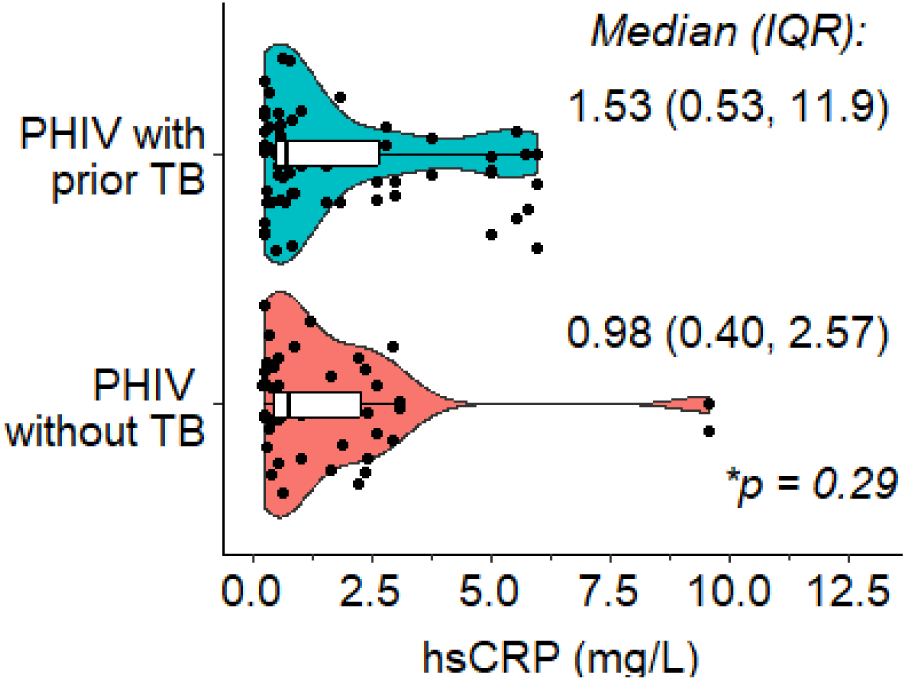
The distribution of high sensitivity C-reactive protein in perinatally HIV-1-infected adolescents according to tuberculosis disease status. * P value for comparison of median hsCRP by TB status.

**Table 2.** Correlates of high-sensitivity C-reactive protein (hsCRP) in perinatally HIV-1-infected adolescents according to tuberculosis disease status.

| Characteristic | <sup>a</sup>Mean difference (95% CI) in median hsCRP (mg/dL) |  |  |  |
| --- | --- | --- | --- | --- |
|  | Unadjusted | P value | Adjusted* | P value |
| No prior TB | *Ref.* |  | *Ref.* |  |
| Prior TB | 0.55 (-1.78, 2.88) | 0.64 | 0.32 (-1.89, 2.53) | 0.78 |
| Male | *Ref.* |  | *Ref.* |  |
| Female | 0.13 (-1.49, 1.75) | 0.87 | -1.03 (-2.94, 0.89) | 0.29 |
| 1-year age increase | 0.44 (0.19, 0.70) | 0.001 | 0.46 (-0.15, 1.06) | 0.14 |
| 2.5 kg/m<sup>2</sup> increase in BMI | 0.56 (0.19, 0.93) | 0.003 | 0.35 (-0.60, 1.30) | 0.47 |
| PI exposure (*vs. none*) | 0.65 (-1.24, 2.540) | 0.49 | 1.05 (-2.07, 4.17) | 0.51 |
| NNRTI exposure (*vs. none*) | 1.31 (-0.04, 2.66) | 0.058 | 1.38 (-1.19, 3.96) | 0.29 |
| Undetectable HIV viral load | *Ref.* |  | *Ref.* |  |
| Viremia | 0.90 (-0.99, 2.79) | 0.35 | 9.78 (0.50, 19.1) | 0.039 |

Values are mean (lower, upper bound 95% CI).
<sup>a</sup> β-coefficients (95% CI) from quantile regression models reported as mean difference in 50<sup>th</sup> percentile (or median) value.
\* Model adjusted for TB history, sex, age, body mass index (BMI), protease inhibitor (PI) and non-nucleoside reverse transcriptase inhibitor (NNRTI) exposure, and HIV viremia.

### Prior TB and left ventricular indices

The differences in cardiac indices by, and their association with TB are summarized in **Tables 3** and **4**. We found no evidence of significant differences in ventricular mass, volumes, and diastolic function according to TB infection. Similarly, there were no significant differences by TB status in scarring [LGE presence: 52.0 vs. 48.4%; p=0.79], diffuse fibrosis [ECV (%): 28.5 vs. 29.1%; p=0.51] or myocardial tissue inflammation [T2 (ms): 38.2 vs. 38.3 ms; p=0.69]. Systolic function measured by LVEF was similar between APHIV with previous TB [mean (95% CI): 63.5 (61.2, 64.6)%] and those without TB [60.6 (58.8, 62.4)%; p=0.048]. This was the case too when systolic function was measured by peak systolic strain. For example, mean (95%CI) peak GLS was −20.7 (−21.6, −19.7)% for APHIV with prior TB and −20.4 (−21.3, −19.5)% (p=0.72) for those without prior TB (**Table 3**).

**Table 3.** Left ventricular parameters in perinatally HIV-1-infected adolescents according to tuberculosis disease status.

| Parameter | Clinical tuberculosis disease status |  |  |
| --- | --- | --- | --- |
|  | No prior TB | Prior TB | P value |
| Number | 27 | 43 |  |
| ***Ventricular-arterial coupling*** |  |  |  |
| Ea (mmHg/mL) | 1.50 (1.39, 1.62) | 1.49 (1.39, 1.60) | 0.93 |
| Height-adjusted Ea (mmHg/mL.m<sup>2.7</sup>) | 0.45 (0.39, 0.51) | 0.46 (0.42, 0.50) | 0.65 |
| Ees (mmHg/mL) | 2.34 (2.15, 2.53) | 2.59 (2.38, 2.80) | 0.039 |
| Height-adjusted Ees (mmHg/mL.m<sup>2.7</sup>) | 0.70 (0.61, 0.78) | 0.80 (0.71, 0.89) | 0.033 |
| Ea/Ees ratio | 0.66 (0.62, 0.70) | 0.56 (0.53, 0.64) | 0.015 |
| ***Ventricular wall strain and function*** |  |  |  |
| Peak circumferential strain |  |  |  |
| Systolic strain (%) | -21.5 (2.3) | -21.3 (2.7) | 0.77 |
| Diastolic strain rate (/s) | 1.76 (1.62, 1.90) | 1.77 (1.64, 1.90) | 0.90 |
| Peak longitudinal strain |  |  |  |
| Systolic strain (%) | -20.4 (-21.3, -19.5) | -20.7 (-21.6, -19.7) | 0.72 |
| Diastolic strain rate (/s) | 1.61 (1.47, 1.74) | 1.63 (1.55, 1.71) | 0.83 |
| LVEF (%) | 60.6 (58.8, 62.4) | 63.5 (61.2, 64.6) | 0.048 |
| ***LV geometry*** |  |  |  |
| LVMi (g/m<sup>2.7</sup>) | 56.8 (53.6, 59.9) | 56.2 (53.4, 57.5) | 0.78 |
| LVEDVi (ml/m<sup>2.7</sup>) | 37.2 (34.8, 39.1) | 36.6 (35.0, 38.9) | 0.71 |
| LVESVi (ml/m<sup>2.7</sup>) | 14.9 (13.5, 15.9) | 13.4 (12.7, 14.9) | 0.088 |
| ***Myocardial tissue*** |  |  |  |
| Presence of LGE | 52.0 (32.7, 70.8) | 48.4 (31.4, 65.8) | 0.79 |
| Extracellular volume (%) | 28.5 (27.5, 29.5) | 29.1 (28.3, 29.9) | 0.51 |
| Native T2 (ms) | 38.2 (37.5, 38.9) | 38.3 (37.4, 39.2) | 0.69 |

Values are reported as mean (95% CI).
Ea = arterial elastance, Ees = end-systolic ventricular elastance; LV = left ventricle; LVM = LV mass; LVEDV = LV end diastolic volume; LVESV = LV end systolic volume.
Postscript (i) = indexed to (height)<sup>2.7</sup>.

### Ventricular-arterial coupling

Regarding our primary endpoint, both adjusted and unadjusted analyses found a lower VAC ratio in PHIV with prior TB than PHIV alone. Prior TB was associated with an adjusted mean difference in Ea/Ees ratio of −0.09 (−0.16, −0.01) (p = 0.048) relative to no prior TB. This difference was driven by the higher Ees in PHIV with TB than PHIV without TB [adjusted mean difference: 0.11 (0.01, 0.22) mmHg/mL.m^2.7^; p=0.031] given the similar Ea [0.01 (−0.07, 0.08) mmHg/mL.m^2.7^; p=0.84]. The other significant determinant of Ea/Ees ratio was sex whereby males had lower Ea/Ees ratio than their female counterparts [adjusted mean difference: −0.09 (−0.15, −0.01); p =0.048].

## Discussion

Our study aimed to assess the impact of TB/HIV comorbidity on CMR-assessed cardiac status in perinatally HIV-1-infected adolescents (APHIV) in South Africa. We found that prior TB versus none was associated with worse cardiac efficiency related to mismatched arterial elastance and ventricular end-systolic elastance. This association was not accounted for by mediated effects of increased hsCRP, a measure of systemic inflammation. Neither did we find significant TB-related differences in ventricular volumes, dimensions, and function. The clinical significance of the observed altered VAC associated with TB requires further studies, as do the underlying immunological mechanisms.

VAC influences cardiac stroke work and cardiac efficiency. Stroke work quantifies the energy expended to generate stroke volume during each cardiac cycle. It is determined by the stroke volume (LVSV) and the mean arterial pressure (MAP). An increase in stroke work [LVSV x MAP] indicates a greater mechanical load on the heart. Cardiac efficiency, on the other hand, is the ratio of stroke work to the total energy expended [stroke work/myocardial oxygen consumption]. It measures cardiac effectiveness in converting energy into useful mechanical work.^22, 23, 29^ Thus mismatched Ea and Ees can result in increased stroke work and decreased cardiac efficiency. In contrast, when coupling is well-matched, stroke work is optimized while cardiac efficiency is increased. Experimental data^23^ suggest that stroke work is maximized with a VAC ratio equals 1 while mechanical and energy efficiency is maximized at a VAC ratio equals 0.5. Nevertheless, in extensive studies involving healthy adult populations, the VAC ratio typically ranged from 0.6 to 0.8.^34, 35, 36^ This observation implies that under normal physiological conditions, the parameters are configured to optimize mechanical and energy efficiency. Indeed, both hypertensive and heart failure patients have been found to have reduced Ea/Ees (<0.6) compared to healthy controls.^36, 37^

We found that in APHIV with prior TB, the VAC ratio was lower compared to APHIV without TB. This would suggest maximization of mechanical and energy efficiency as opposed to stroke work.^23^ However, the VAC ratio with TB/HIV comorbidity [Ea/Ees = 0.56] may represent cardiovascular efficiency below the optimal range.^37, 38^ Low values of the VAC ratio imply inappropriately high ventricular end-systolic elastance for a particular level of arterial elastance. In our study, arterial elastance was equal between the two TB groups whereas ventricular end-systolic elastance was higher among those with TB than their counterparts without. There are at least two mechanisms by which a low VAC ratio might portend adverse cardiovascular outcomes. It has been shown in adults that a high Ees increases the cardiac energy cost of increasing stroke volume^37, 38^, and that low VAC ratio is associated with increased diastolic stiffness and diastolic dysfunction.^37^

Our findings corroborate the evidence from the few available studies showing an association between TB/HIV comorbidity and heart failure or its antecedents. Bakari *et al.,*(2013) found that a history of TB was associated with a 3-fold higher likelihood of LV systolic heart failure [adjusted OR: 3.01 (1.32 - 11.56)] among adult persons living with HIV (PLWH) in Tanzania^39^, whereas Ndongala *et al.,* (2022) in Lesotho reported a 6-fold higher likelihood of heart failure [adjusted OR: 6.25 (1.24 - 31.48)] with prior TB.^40^ In the latter, their definition of heart failure included pulmonary heart disease, i.e., right heart failure in the presence of pulmonary hypertension. Similarly, past TB was independently predictive of subclinical cardiopulmonary dysfunction [adjusted OR: 2.3 (1.2 to 4.4)] in South African adolescents with PHIV.^41^ This is the only study to date, to our knowledge, focusing on APHIV. Noteworthy, this study defined cardiopulmonary dysfunction as any of RV systolic dysfunction, LV diastolic or systolic dysfunction or abnormal mean pulmonary arterial pressure, in conjunction with abnormal spirometry or a deficient 6-minute walking test. Detracting from these studies, including ours, is their small sample size, cross-sectional design, convenient sampling, and hospital- or health-facility based enrolment.

The differences by TB status in systemic inflammation were not statistically significant, and thus precluded mediation causal analyses. Our study may have been underpowered to detect differences in levels of hsCRP. It remains an important and urgent priority, from an intervention point of view, to delineate the role of immune activation and systemic inflammation in TB/HIV associated cardiovascular changes. Relatedly, assessing Ea and Ees over traditional indices like LVEF has the advantage of improved discrimination of changes in ventricular performance, arterial load or both.^22^ Formally evaluating the utility of this approach to risk identification and stratification in APHIV will be an important extension of our work. However, this presupposes that our observed and otherwise subclinical findings of mismatched arterial elastance and ventricular end-systolic elastance have prognostic significance.

## Strengths and limitations

We believe that our study is among the first to examine the relationship between HIV/TB comorbidity and VAC in APHIV. This is an understudied but potentially at-high risk population subgroup for premature cardiovascular ill-health. We employed CMR which has superior reproducibility, higher accuracy and sensitivity for cardiac assessment compared to other imaging modalities. Further, CMR-based assessment of VAC parameters has been demonstrated to provide estimates that are comparable to those derived from invasively measured intracardiac pressure–volume loops.^24^

VAC can detect subtle changes in cardiovascular function at earlier stages compared to traditional indices like LVEF which may mask heart dysfunction until it is well advanced.^42^ However, we lacked detailed TB infection history. Neither did we comprehensively assess immune activation and systemic inflammatory pathways. These shortcomings precluded granular mechanistic insights. Besides, our study may have been underpowered to detect differences in hsCRP, and thus to undertake a causal mediation analysis. Equally cautionary was our definition of TB which included clinical diagnoses and thus probably misclassification biases. Notwithstanding, our findings do shed new light on a subject of growing clinical and public health concern. These findings remain to be replicated, and their long term significance mapped out. Future work should include HIV uninfected adolescents as controls to better assess and understand the impact of TB/HIV comorbidity on cardiovascular health.

## Conclusion

We demonstrate that previous TB in APHIV is associated with suboptimal cardiac efficiency, related to mismatched arterial elastance and ventricular end-systolic elastance. The clinical significance of these findings requires further studies, including a wider range of biomarkers of specific immune pathways. However, it is clear that greater efforts are needed for enhanced and effective interventions to optimize the management of APHIV particularly as they enter adulthood, and become increasingly independent of their caregivers.

## Data Availability

All data produced in the present study are available upon reasonable request to the authors..

## Acknowledgements

In addition to thanking all study participants and their families, and the HBNU Fogarty Global Health Training Program, the authors would also like to acknowledge the support of the following people who made the study possible: Sana Mahtab, Nana Akua Asafu-Agyei, Landiwe Daka, Sharon Wakefield, Shanaaz Davids, Nikelwa Mvango-Njemla, Nomawethu Jele, Nomandla Udogwu*, Pumeza Nazo, Njemia Nikelwa, Keyola George, Akhona Mazingi, Tafadzwa Mautsa, Mothabisi Nyathi, Faith Qeja, Luyanda Nkondlwana, Mazwi Maishi, Mariaan Jaftha, Daniel Doetz, Liezl Julius, Michael Wheeler, Shaheed Sorathia, Nabeal Kaskar, Dave Nshuntishema, Okeyo, Patricie Niyitegeka, and Ben Jann.

*posthumously.

## Conflicts of interest

The authors have no conflicts of interest to disclose.

---

**Supplementary Figure 1.**
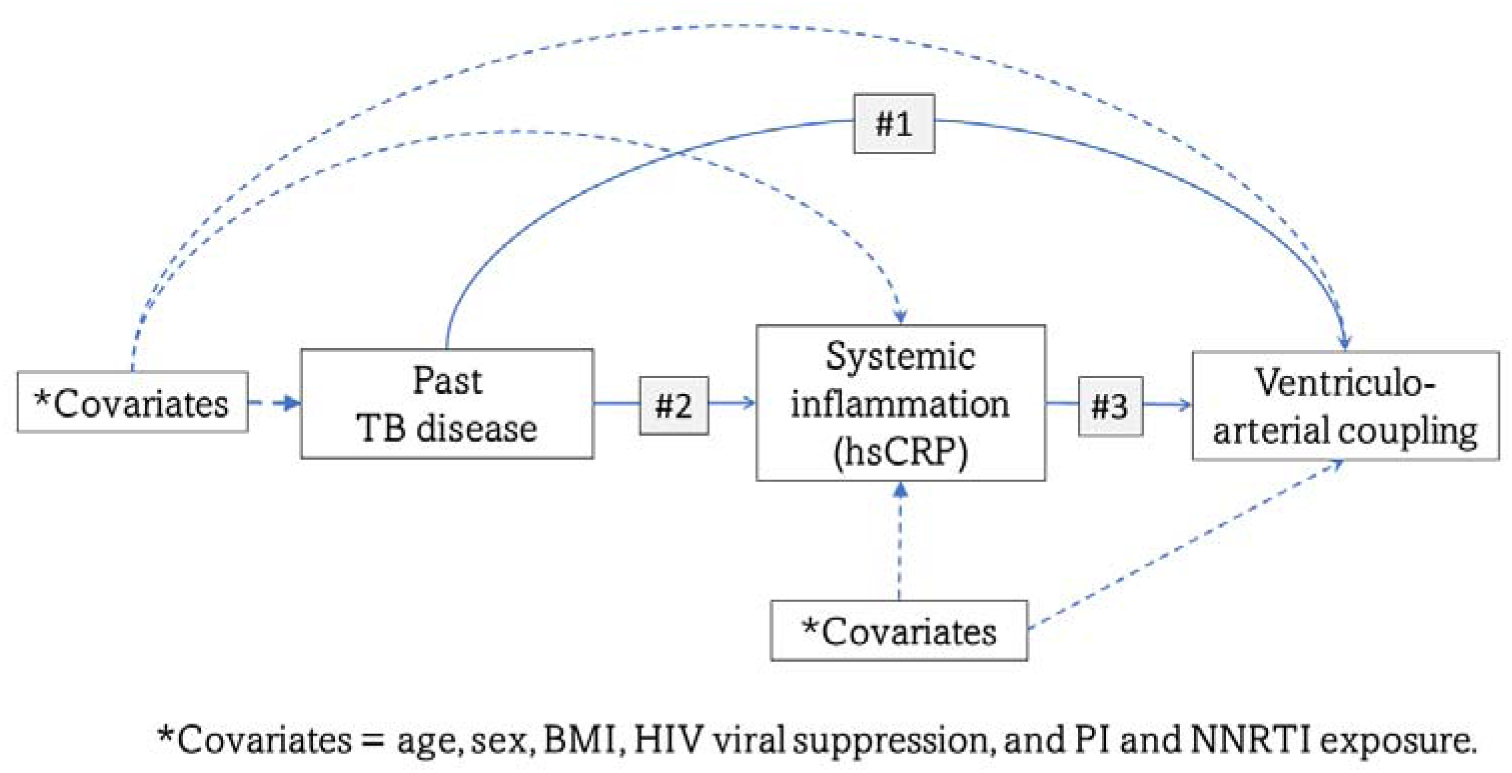
Mediation models demonstrating indirect effect of inflammation in the relationship between past tuberculosis disease and ventriculo-arterial coupling in perinatally HIV-1-infected adolescents. #1. Linear regression model: VAC = α_0_+ α_1_.TB + α_2_.Covariates #2. Quantile regression model: hsCRP = β_0_ + β_1_.TB + β_2_.Covariates #3. Linear regression model: VAC = δ_0_+ δ_1_.hsCR + δ_2_.Covariates #4. Linear regression model: VAC = γ_0_ + γ_1_.TB + γ_2_.hsCR + γ_3_.Covariates

## Notes

**Support** Itai M. Magodoro was supported by a training award from the Fogarty International Center and National Institute of Mental Health of the National Institutes of Health (D43 TW010543) and the AIDS Healthcare Foundation, Los Angeles, USA. Heather Zar is supported by the South African Medical Research Council (SA MRC). CTAAC was funded by the NIH (R01HD074051; PI Heather Zar). Ntobeko Ntusi gratefully acknowledges funding from the National Research Foundation, South African Medical Research Council, US National Institutes of Health, Medical Research Council (UK), and the Lily and Ernst Hausmann Trust.

### Competing Interest Statement

The authors have declared no competing interest.

### Funding Statement

Itai M. Magodoro was supported by a training award from the Fogarty International Center and National Institute of Mental Health of the National Institutes of Health (D43 TW010543) and the AIDS Healthcare Foundation, Los Angeles, USA. Heather Zar is supported by the South African Medical Research Council (SA MRC). CTAAC was funded by the NIH (R01HD074051; PI Heather Zar). Ntobeko Ntusi gratefully acknowledges funding from the National Research Foundation, South African Medical Research Council, US National Institutes of Health, Medical Research Council (UK), and the Lily and Ernst Hausmann Trust.

### Author Declarations

The Human Research Ethics Committee (HREC) of the Faculty of Health Sciences of the University of Cape Town (HREC 051-2013) approved all study activities.

